# A strengths-based, participatory qualitative exploration of digital information access and critical health literacy among culturally and linguistically diverse communities in Australia

**DOI:** 10.64898/2026.06.24.26356477

**Authors:** Danielle M Muscat, Nada Mustapha-Khodragha, Melanie Wong, Suci Trisnasari, Gayatri Talla, Fika Rizkyanti, Sharon Ng, Gauri Kapoor, Saad Khalid, Lina Rigby, Naglaa Sourour, Belinda Peri, Thi Mai Nguyen, Olivia Mac, Hooi Min Lim, Kirsten McCaffery, Julie Ayre

## Abstract

As the volume of online health information expands, critical health literacy is increasingly positioned as a key resource for navigating the contemporary “infodemic”. This qualitative study examines how multi-lingual adults from diverse migrant backgrounds in Australia access and appraise online health information. Grounded in a participatory action research approach, bilingual community co-researchers (n=11), collectively speaking eight languages other than English, were embedded across all stages of the research. Semi-structured interviews with 55 participants were conducted in participants’ preferred languages. Underpinned by a critical realist epistemology and a strengths-based approach, we undertook an iterative, inductive Framework analysis. We constructed two themes (comprising 7 subthemes): (1) Navigating “a jungle” and (2) “It’s always a mix of trust and scepticism”. Our findings suggest that culturally and linguistically diverse communities are required to navigate demanding information environments characterised by an overwhelming volume of online health content, much of it not tailored to diverse language needs or literacy levels and drawn from sources that are often difficult to verify. Notwithstanding these structural challenges, our strengths-based analysis identified a repertoire of practices through which participants worked to locate understandable information and distinguish credible sources from misinformation. AI tools emerged as a resource in this process, operating as an extension of participant’s own cognitive and linguistic work to simplify and synthesise complex information, but positioned ambivalently within credibility practices. These findings have implications for the development of culturally responsive health literacy resources that build on the existing capacities of culturally and linguistically diverse communities.

## 1. Introduction

Never in human history has there been such an abundance of health information available. Although the internet, social media and emerging artificial intelligence (AI) platforms offer unprecedented opportunities for the dissemination of evidence-based health information (1, 2), limited governance of many online environments has intensified challenges related to the spread of misinformation (inaccurate information) and disinformation (knowingly false information intentionally designed to mislead) (3–5). This evolving information ecosystem has contributed to an increasingly complex health information landscape in which individuals must navigate vast volumes of content of uneven credibility (4).

Within this context, attention has increasingly turned to the concept of critical health literacy and its role in enabling individuals and communities to engage meaningfully with health information (6–8). Nutbeam conceptualised critical health literacy as the advanced cognitive and social skills required to critically appraise, contextualise and apply health information in ways that enhance individual and collective control over health-related decisions (9). Beyond functional access to information and abilities related to reading and understanding text, critical health literacy encompasses the ability to evaluate the reliability, validity, credibility and applicability of sources and interpret health messages within broader social, cultural, and political contexts (9, 10). Here, critical health literacy is seen to reflect *“the individual’s capacity to contextualise health knowledge for his or her own good health, to decide on a certain action after a full appraisal of what that specific action means for them “in their own world””* (11). As the volume of online health information continues to expand, critical health literacy has been identified as a key protective factor against the harms associated with the contemporary “infodemic” (8, 12).

Alongside a growing body of conceptual and definitional work related to critical health literacy (e.g. (10, 13, 14)), there have been repeated calls to embed critical health literacy competencies within school, university, and community education curricula (15). In parallel, a range of educational interventions have been developed to strengthen these competencies. Early conceptualisations, particularly those advanced by Steckelberg and colleagues, framed critical health literacy largely as competence in the principles of evidence-based medicine, encompassing knowledge of medical concepts, research design, basic statistics, and the skills required to locate, interpret, and critically appraise scientific evidence (16). More recent initiatives have expanded this perspective. Programs such as Informed Health Choices (17, 18) place greater emphasis on the appraisal of health information and claims encountered in everyday life, including through social media, news reporting, advertising, and other digital platforms. These approaches focus on developing skills to recognise unreliable claims, assess the credibility of sources, identify common reasoning errors, and judge whether available evidence provides a trustworthy basis for action.

While these interventions provide structured approaches to developing critical health literacy competencies, little is known about how critical health literacy is enacted in everyday life within increasingly complex digital health environments. Research to date has also been concentrated in Western contexts, particularly Europe, North America, Australia, and New Zealand, with limited attention beyond these countries or to culturally and linguistically diverse communities (19). In contrast, a broader body of research has examined susceptibility to misinformation in these communities, often adopting a deficit-oriented framing. These studies frequently emphasise heightened vulnerability to misinformation among culturally and linguistically diverse communities due to greater reliance on social media and community networks (20–22), language barriers (20, 22–26), lower health literacy (20, 23, 27–29), and challenges navigating increasingly complex digital health systems (20). For example, work during the COVID-19 pandemic found that individuals from migrant backgrounds were 11 times more likely to endorse unsubstantiated COVID-19 prevention claims compared with those with high English proficiency (30). While these findings highlight important inequities, an exclusive focus on deficits risks obscuring the existing strengths, resources and adaptive strategies within culturally and linguistically diverse communities which support the navigation and appraisal of health information. Strengths-based approaches to health literacy instead foreground factors that support health-related decision-making, including individual assets and community-level resources (31).

Yosso’s (2006) concept of “community cultural wealth” exemplifies a broader shift towards strengths-based and asset-oriented approaches that move away from deficit framings of culturally and linguistically diverse communities. Rather than focusing on perceived gaps or limitations, this perspective foregrounds the diverse cultural knowledge, skills, abilities, and networks that are often unrecognised or undervalued in dominant systems (32). Drawing on critical race theory, Yosso articulated multiple forms of capital embedded within community cultural wealth, including aspirational, navigational, social, linguistic, familial, and resistant capital, which are defined in Box 1. Originally developed in education research, Yosso’s work has contributed to a broader shift in public health and health promotion towards strengths-based approaches that foreground how culturally and linguistically diverse communities draw on existing forms of knowledge, skills, and networks to navigate structural barriers and support health-related decision-making. While this perspective has informed a growing body of scholarship, it has not yet been explicitly integrated with critical health literacy.

#### Box 1. Forms of capital from Yosso’s (2006) model of community cultural wealth

1. *Aspirational capital* - the ability to maintain hopes and dreams for the future, even in the face of real and perceived barriers.
2. *Linguistic capital* - the intellectual and social skills attained through communication experiences in more than one language and/or style.
3. *Familial capital* - cultural knowledges nurtured among *familia* (kin) that carry a sense of community history, memory and cultural intuition.
4. *Social capital* - networks of people and community resources that can provide both instrumental and emotional support to navigate through society’s institutions.
5. *Navigational capital* - skills of manoeuvring through social institutions.
6. *Resistant capital -* knowledges and skills fostered through oppositional behaviour that challenges inequality.

Adopting a strengths-based lens, this study examines the individual, social, and cultural resources that support critical health literacy within culturally and linguistically diverse communities. In the context of increasingly complex digital health environments, characterised by an abundance of health information, misinformation, competing claims, and diverse sources of varying credibility, we explore how individuals draw on these resources to navigate, evaluate, and respond to health information encountered online.

## 2. Methods

### Study design

This study employed a participatory action research (PAR) qualitative design, grounded in research traditions that emphasise co-production of knowledge, community ownership and action-oriented inquiry (33). PAR was selected to address structural inequities in research participation and knowledge production by placing the expertise of community members from diverse cultural and language backgrounds at the centre of this study. The study was conducted between 2021 to 2025 in Australia, with ethics approval from the University of Sydney Human Research Ethics Committee (2021/244). All participants gave informed consent prior to commencement of interviews.

Reporting of this manuscript has been guided by the Big Q Qualitative Reporting Guidelines (BQQRG) (34).

### Community researcher engagement and training

Consistent with PAR principles, community members were engaged as bi-lingual community co-researchers throughout all stages of the research process, including study design refinement, participant recruitment, data collection, analysis, interpretation and dissemination. Academic researchers functioned as facilitators and methodological supports rather than sole knowledge producers.

Bi-lingual community co-researchers (n=11) were engaged through existing consumer groups (the Sydney Health Literacy Lab Community panel (Co-SHeLL) (35) and Western Sydney Local Health District Consumer Council (36)), as well as targeted email communication circulated through trusted organisational channels to reach bi-lingual community members positioned to serve as cultural and linguistic brokers. All co-researchers were fluent in English and at least one other language, had established trust and strong community connections, and demonstrated an interest in research and community advocacy.

Community researchers completed structured training in qualitative research methods, with ongoing mentorship throughout the study to support methodological rigor and researcher wellbeing. Combined, community researchers spoke 8 languages other than English: Arabic, Bahasa Indonesia, French, Hindi, Mandarin, Punjabi and Vietnamese.

### Participant eligibility and recruitment

Eligible participants were adults aged over 18 years who spoke a language other than English at home. They were recruited using community-based, culturally responsive strategies, including peer referral, outreach through community organisations, and in-language recruitment materials. Recruitment was conducted primarily by community co-researchers, leveraging existing relationships to enhance trust and accessibility. To minimise the risk of coercion, our recruitment approach emphasised that participation was voluntary and that declining participation would not affect existing community relationships or access to services, and informed consent was obtained independently of recruitment activities.

Our inclusion criteria were intentionally broad to allow for diversity in gender, age, education, specific language spoken at home, time spent living in Australia and level of health literacy, with purposive sampling adopted to try to promote variation in these demographic factors where possible. Sampling decisions were revisited iteratively throughout data collection through regular review of participant characteristics and preliminary analyses, enabling responsive refinement of recruitment efforts.

### Data collection

Data were collected through semi-structured, in-depth interviews conducted in participants’ preferred language. Semi-structured interviews were chosen for their flexibility, allowing in-depth exploration of participants’ experiences while maintaining a consistent framework across interviews (37). Interviews were conducted by trained bi-lingual community researchers, either in person or remotely (via Zoom) depending on participant preference and public health considerations. Prior to data collection, some community researchers were known to participants through existing relationships within community organisations, supporting trust and engagement but also requiring careful attention to voluntary participation, confidentiality, and transparent disclosure of the research role and boundaries. During interviews, researchers adopted a reflexive stance, explicitly acknowledging their dual role as both community members (insider) and researchers, and using this positionality to support rapport while being mindful of potential power imbalances. Training emphasised maintaining neutrality, avoiding leading questions, and minimising the influence of researchers’ views or prior relationships on participants’ responses.

The interview guide was co-developed by academic and community researchers, piloted prior to data collection, and iteratively revised throughout the study to explore emerging lines of enquiry. Consistent with a participant-centred approach, participants were invited to nominate a health topic of their own choosing and to reflect on their experiences of seeking and engaging with online health information related to that self-defined concern. Terminology such as “health literacy” and “critical health literacy” was deliberately not used during interviews to avoid imposing researcher-defined constructs on participants’ accounts. Demographic information collected at the end of the interview included: age, gender, education attainment, country of birth, language(s) spoken at home and health literacy (measured using a validated single item screener (38)).

Interviews lasted approximately 20-45 minutes, were audio-recorded with consent, and transcribed and translated into English by community researchers. All participants that completed the interview received a $25 gift card.

### Sample size

Rather than defining sample adequacy through numerical saturation, this study drew on the non-positivist qualitative concept of ‘information power’ (39), with emphasis placed on the richness, relevance, and analytic depth of the dataset in relation to the research aims and questions. Dataset adequacy was continuously assessed throughout data collection and analysis through iterative, team-based reflexive discussions. We explicitly considered two dimensions that strengthened information power within this study: (i) the quality and depth of dialogue, supported through in-language interviews and culturally responsive interviewing practices, and (ii) the strong theoretical anchoring in critical health literacy. These factors were weighed against dimensions that increased the required sample size to achieve sufficient analytic depth: (iii) the breadth of the study aims, (iv) the heterogeneity of the sample, including cultural and linguistic diversity, demographic variation, and differences in online health information-seeking experiences; and (v) the progressive development of cross-case analytic insight, which necessitated a larger dataset to adequately capture variation and refine emerging interpretations.

### Data Analysis

Aligned with a critical realist epistemology, this study assumes that knowledge is both contextual and fallible, shaped by social, cultural, and institutional conditions, while also referring to a reality that exists independently of our perceptions (40). In this sense, findings are understood as partial representations of underlying mechanisms, co-constructed through the interaction between participant accounts and researcher interpretation. Reflexivity was therefore integral throughout the analytic process, with attention to how researcher positioning and theoretical framing shaped interpretation.

Data were analysed using Framework Analysis, following the approach outlined by Gale et al. (41). Framework analysis offers a structured yet flexible method well suited to applied qualitative research, enabling systematic engagement with large and complex datasets while retaining close proximity to the raw data (40). The stages of Framework Analysis support transparency and rigour by ensuring that data can be systematically retrieved, compared, and interrogated at multiple points throughout the analytic process (42).

In this study, Framework analysis was used in a way consistent with a critical realist orientation, accommodating both inductive and deductive forms of reasoning and a combination of data driven, researcher driven, and theory driven processes. Inductive coding during early stages allowed for the generation of meaning grounded in participants’ accounts, while later stages of charting and mapping supported more structured comparison across cases and the development of higher-order interpretive explanations. In this way, Framework Analysis functioned as a transparent and auditable scaffold for progressing from descriptive accounts to more explanatory interpretations consistent with a critical realist approach (40).

Stages of Framework analysis as applied in this study are detailed below. While presented sequentially for clarity, the analytic process was iterative and recursive in practice, with ongoing movement back and forth between stages. This allowed continual refinement of coding, categories, and interpretations as analysis progressed, rather than proceeding in a strictly linear fashion. The analysis process was supported by weekly team meetings involving all academic and community co-researchers.

1. **Familiarisation** – All transcripts were read multiple times by both community and academic researchers, with co-researchers also encouraged to listen to all or selected portions of the audio recordings where useful. During team meetings, we discussed any contextual and reflective notes recorded during the interviews, alongside overall thoughts and impressions throughout the familiarisation phase.
2. **Coding –** Each transcript was examined line by line and assigned codes using an open coding approach, whereby all potentially relevant content was coded. Academic researchers (DMM and NMK) independently coded the initial two transcripts for each co-researcher to support consistency and shared interpretive development.
3. **Developing a working analytical framework** – After coding the initial transcripts, all researchers met to compare assigned labels and to develop a shared set of codes for application to subsequent transcripts. This iterative process involved several meetings to refine and consolidate the coding approach. Codes were then organised into higher-order categories, which were defined by the academic research team with an emphasis on plain language and conceptual clarity. Together, these categories constituted a working analytical framework.
4. **Applying the analytical framework** – All members of the research team applied the analytical framework by indexing subsequent transcripts using the existing categories and codes.
5. **Charting data into the framework matrix** – An Excel spreadsheet was used to construct a matrix into which data were charted, summarising findings by category for each transcript. The team met regularly to consider how best to balance data reduction with the need to retain the original meaning and “feel” of participants’ accounts and this was supported through the inclusion of illustrative and analytically significant quotes within the matrix. Charting undertaken by community co-researchers was independently checked by the academic team to ensure consistency and accuracy.
6. **Interpreting the data** – Data interpretation was iterative, with themes and sub-themes analysed to identify patterns, relationships, and explanatory constructs. DM prepared analytic memos which were discussed and interrogated within the broader research team during team meetings, drawing on both participant accounts and contextual understanding to move beyond description towards explanation in line with a critical realist analytic logic.

### Reflexivity Statement

Understanding that meaning in qualitative research is co-constructed by participants and researchers, our team engaged in reflexivity as an active, ongoing process of critical self-reflection about how our positioning as academics and community co-researchers shaped research questions, influenced data generation and informed analytic decisions and interpretations. While reflexivity permeated all stages of the study, this section foregrounds two key considerations: the role of community co-researchers and the use of a strengths-based analytic approach.

Interviews were conducted by community co-researchers in participant’s preferred language. This approach supported rapport, cultural safety and participants’ ability to express nuanced meanings that may not have emerged if all interviews were conducted in English. Co-researcher’s shared linguistic and cultural background positioned them as an ‘insiders’, shaping both the interview dynamics and the knowledge produced. At the same time, this insider position required ongoing reflexive attention; the co-researcher’s own embeddedness in local norms also shaped questioning, probing and interpretation, requiring active reflection to avoid over-familiarity and unexamined assumptions. Regular reflexive discussions and debriefing sessions were held to identify potential biases in participant recruitment, data collection, interpretation and analysis. These sessions facilitated recognition of how social, cultural, and linguistic backgrounds shaped the framing of questions, the interpretation of narratives and the prioritisation of emergent themes.

We purposefully approached the analysis with a strengths-based orientation. This positioning shaped attention toward participants’ capacities, resources, and adaptive strategies and accounts of agency and resourcefulness, influencing both coding and theme development. Reflexive engagement was maintained throughout analysis through memo-writing and iterative questioning of analytic decisions, particularly where an emphasis on strengths risked obscuring experiences of structural inequity, complexity or distress. The goal here was to honour participants’ complex and contextualised accounts by ensuring that attention to strengths did not eclipse the conditions shaping their experiences but instead remained grounded in participants’ own meaning-making.

## 3. Results

### Demographics

Demographic characteristics of participants are shown in Table 1 (N=55). A majority of participants identified as female (75%), with most aged 18–44 years (69%). Participants were born in 17 countries outside of Australia, most commonly within the Southeast Asian (35%) and Indian Subcontinent (31%) regions. Twenty-three participants (41.8%) had lived in Australia for less than five years.

**Table 1.** Participant demographic characteristics (N=55)

| Characteristic | n (%) | Characteristic | n (%) |
| --- | --- | --- | --- |
| Gender |  | Self-reported English proficiency |  |
| Female/woman | 41 (75) | Very well | 11 (20) |
| Male/man | 13 (24) | Well | 30 (55) |
| Non-binary | 0 (0) | Not well | 3 (5) |
| Prefer not to say | 1 (2) | Not well at all | 1 (2) |
|  |  | Missing | 10 (18) |
| Age (years) |  | Health literacy |  |
| 18-24 | 10 (18) | Adequate | 38 (69) |
| 25-34 | 13 (24) | Limited | 17 (31) |
| 35-44 | 15 (27) |  |  |
| 45-54 | 9 (16) |  |  |
| 55-64 | 1 (2) |  |  |
| 65+ | 7 (13) |  |  |
| Years in Australia |  | Region of Birth |  |
| <1 | 5(11) | East Asia | 10 (18) |
| 1-2 | 7(16) | Indian Subcontinent | 17 (31) |
| 3-5 | 11 (20) | Middle East | 4 (7) |
| 6-10 | 11 (20) | Southeast Asia | 19 (35) |
| >10 | 18 (33) | Other | 3 (5) |
| Missing | 3 (5) | Missing | 2 (4) |
| Highest level of education completed |  | Language other than English spoken at home<sup>†</sup> |  |
| Up to year 10 | 4 (7) | Arabic | 6 (11) |
| Yr 12 or equivalent | 10 (18) | Chinese (Cantonese, Mandarin) | 9 (16) |
| Certificate III/IV or diploma | 7 (13) | Indonesian | 10 (18) |
| Bachelor's degree* | 25 (45) | Nepali | 4 (7) |
| Post-graduate degree* | 9 (16) | Tamil | 5 (9) |
|  |  | Urdu | 5 (9) |
|  |  | Vietnamese | 5 (9) |
|  |  | Other | 12 (22) |
| *University Degrees: 5 completed in Australia, 14 completed overseas |  |  |  |
| <sup>†</sup> Participants spoke more than one language |  |  |  |

### Theme overview and conceptual map

Through our analysis, we constructed two themes, comprising seven sub-themes as shown in Figure 1. A third theme relating to familial and social capital was also identified, referring to the ways in which family members and broader social and community networks shaped participants’ critical appraisal of health information. Within this theme, participants drew on interpersonal relationships for guidance, validation, and interpretation of health information, highlighting the relational and collective dimensions of critical health literacy. It is reported separately due to its conceptual richness and distinct analytical focus.

**Figure 1.**
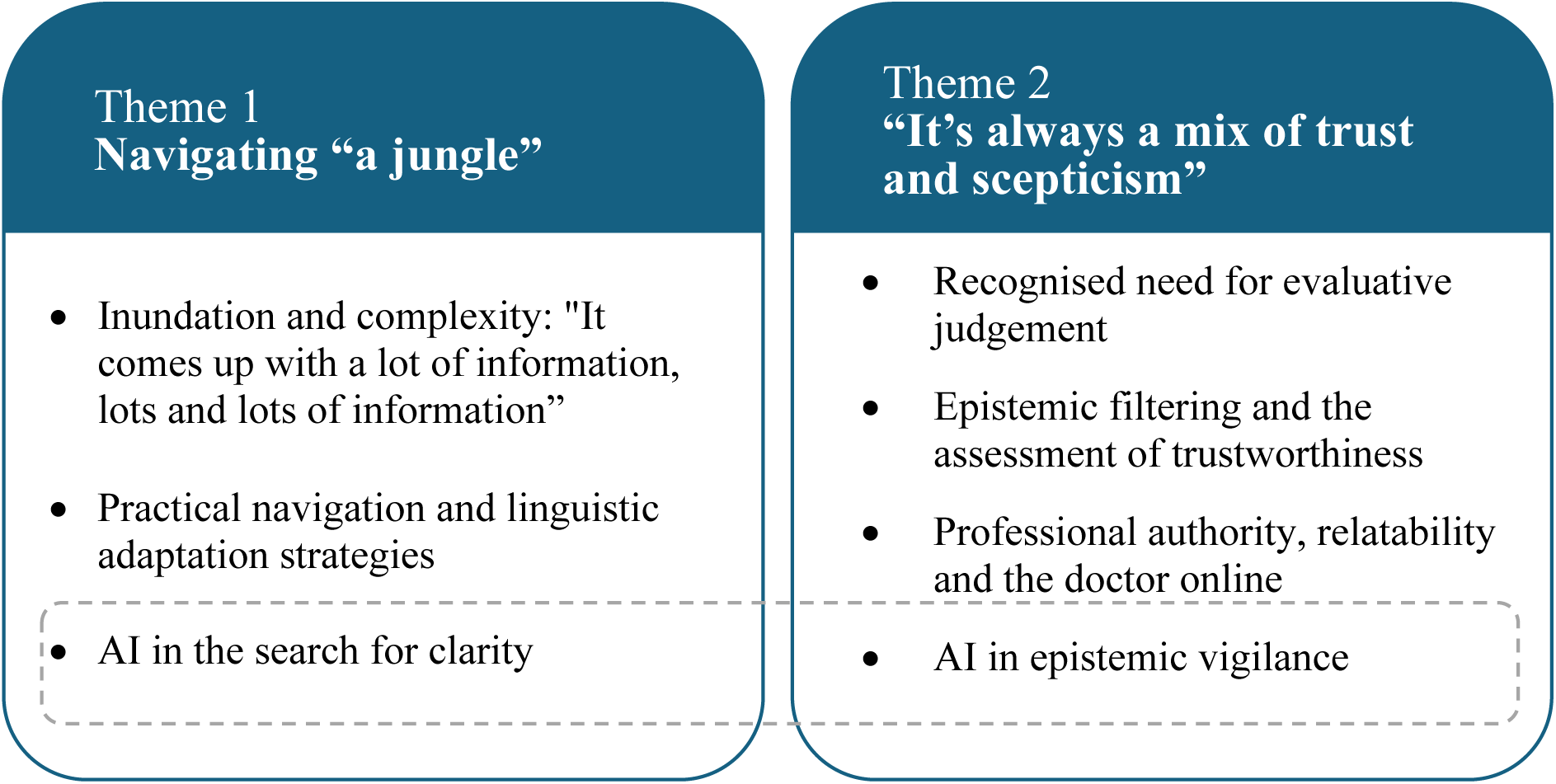
Thematic map.

### Theme 1: Navigating “a jungle”

#### Inundation and complexity: “It comes up with a lot of information, lots and lots of information”

Participants’ accounts depicted online information-seeking as an effortful and often overwhelming process, characterised by exposure to an excess of information, with “*so many links”* (P01) and “*too many sources”* (P03). The abundance of available online information frequently required repeated searches and sustained interpretive labour to discern relevance amongst *“a jungle”* (P49) of information.

> *“So, I’d search on Google, but there’s just so much information—it can be overwhelming”.* P30
>
> “…[I had to] *Google it six times, because too many* [sic] *information…I was confused”.* P07
>
> *“It comes up with a lot of information, lots and lots of information, very confusing. And…lots of links….When I go on Google and say ‘acid reflux’ it comes up with a lot of things, you know, like some of them may not be related to what I’m looking at”.* P52

Participants also described encountering texts they perceived as excessively detailed, contributing to a sense of informational saturation in which quantity of information supplanted clarity. The organisation of information on webpages and apps was frequently experienced as difficult to navigate, requiring users to *“click and click and click”* (P06) through multiple layers of extraneous content that was perceived to obscure rather than clarify key information.

> *“Some articles have a ton of detail, and I end up not understanding what they’re trying to explain—or they just dump too much data”.* P26
>
> *“The second is that some websites are confusing or not to the point—they start with a lot of fluff and don’t get straight to the facts. That’s pretty much it when it comes to health information.”* P30

This navigational burden was compounded by “*lots of jargons* [sic]” (P03) and “*complex scientific explanation*[s]” (P09). For some, but not all, language was seen as a key mediating factor in their comprehension of health information, noting that health information in the English language was particularly “*hard to understand”* (P41).

> *“Obviously, sometimes we…can’t understand the exact meaning…yes, sometimes we misunderstood the information”.* P12
>
> *“Even though I’m fluent in speaking English, some medical terms are hard to understand. For example, bladder-related issues—I know the general area, but not the complications. I need to look up more to understand about the meaning of what I read online”.* P41

Here, access to information was not synonymous with accessibility. The cognitive, interpretive and navigational work demanded by contemporary digital information environments constrained participants’ ability to mobilise information, leaving some feeling *“quite stuck”* (P06).

### Practical navigation and linguistic adaptation strategies

Inundated with complex information, participants described their active efforts to filter information for clarity. Here, self-filtering can be seen as an expression of agency and marker of navigational capital, actively shaping how they accessed, interpreted and used health information within complex digital environments that were not necessarily designed for their success. For example, participants reported prioritising information that they found to be clear and concise, *“where people explain things using everyday language* [and] *more relatable words rather than heavy medical jargon”* (P45). This included content organised into “*informative and bite-size chunks*” (P06) and using “*laymen terms*” (P06), with participants expressing that they *“think it’s better if the information is short and specific to the topic.”* (P30)

> *“I need the information, and I look for something that’s not complicated…”* P28
>
> *“Okay, well, the one that I actually like using…it’s written in lay language that I can easily explain to someone. And it had, the diagrams and all that. So it’s not just words that someone’s just put there… because they’ve actually went to the effort of doing the diagrams. You can say it, you know, you understand it. You can explain it to someone by yeah, pointing to it.”* P49

To this end, participants discussed how they tried to simplify their search terms and/or pro-actively selected platforms that they felt aligned with their informational needs, often prioritising more accessible formats, including *“illustrations”* (P46), image-based results or visual summaries.

> “*So I started it off by going on Google and searching just by the two words, acid reflux….I think that you keep your words very simple. Like, don’t put in too many words in your search; it comes up with a lot of information”.* P52
>
> *If I want something short but clearly explained through speech, I use TikTok. But if I want to understand something by reading, I use Google.* P23
>
> *“When I searched, I saw some research articles from Harvard. Or from places like Stanford. And usually, they’re written quite…academically….So instead, I started searching through Google Images.”* P44

Others described sustained and often labour-intensive strategies to make sense of complex information.

> *“Well, what I have to actually do is I write the word down. And I write everything that I didn’t understand, and then I have to go back, yeah, into a different place, and then, like, still on Google Bar and find out what each word means.”* P49
>
> *“And because I read based on…credibility, so when I see something I trust, I choose to read it. Even if it’s hard to understand, I’ll try to read it thoroughly and understand it completely”.* P44

Once identified, participants emphasised the importance of saving or bookmarking information to avoid repeating the search process.

> *“So when you finally find the information, if you don’t bookmark it or lose track of it, you’ll have to go through the whole process again to find it”.* P27

First-language searching was also reported in participants’ efforts to reduce the cognitive burden of navigating unfamiliar English-language medical terminology. These searches often yielded content considered more accessible and meaningful, frequently sourced from overseas websites.

> *“I search in Vietnamese because I’m not very familiar with medical terminology in English. I feel more confident using Vietnamese”.* P45
>
> *“I first searched in English, but it was hard to understand. Later, I searched in Chinese and found more helpful info —mostly from Taiwanese websites. Australian sites were harder to understand, especially since I’m not in the medical field… I prefer sources that use Chinese because they’re easier for me to understand and comprehend”.* P41
>
> *“… in English sometimes, because I’m looking for some words, maybe new vocabulary. And after that, I change it to Arabic to understand all the situations.”* P53

However, platform algorithms did not always respond as expected. Several participants described experiencing a disconnect between their in-language search intentions and the content delivered to them, producing forms of digital dislocation for migrants seeking culturally and linguistically relevant health information.

> *“So, when I arrived here and searched for things about Indonesia or in my own language, it was kind of hard because the app didn’t understand. It was like everything had switched to Australia, and they didn’t understand Indonesian…”* P23
>
> *“You could switch to Chinese, but the information was cut in half or not updated in real time…The English version had way more content than the Chinese one.”* P43

### AI in the search for cl<u>a</u>rity

In the search for clear information, participants’ engagement with AI reflected a spectrum of active and passive strategies. On the more passive end, many expressed a preference for AI-generated summaries or the “*top result*” (P22) provided by search engines, perceiving these outputs as convenient starting points that offered a general overview of a topic without requiring extensive navigation or evaluation.

> *“Well, to make it easier, I usually read the top result first because it’s the most general. It’s not a website—it’s more like general information from the search engine. Then, below that, actual websites appear. So I read the general info first, which I think is generated by Google, and then I look at the websites below”.* P22
>
> *“Google is* [sic] *just gives you the information exactly at the top”.* P08
>
> *“On Google, I just ask directly. For example, “flu medicine for children.” Just like that, and thankfully I get results right away. But I think it’s AI that answers, right? Because when we search, there’s that top answer, and below that there are accounts like Alodokter, Halodoc, and so on. I usually just look at the top one, because if I open Alodokter or Halodoc, the answers are all different and confusing”.* P28
>
> *“I just describe the issue and get results immediately. It’s like magic—type a little and everything comes out.* P43

Others described more active, deliberate use of AI tools, such as ChatGPT, to summarise lengthy articles or simplify complex explanations. In these instances, participants tailored queries to their informational needs, using the technology as an extension of their own cognitive and linguistic work; they not only relied on the outputs of AI but shaped their use according to their personal comprehension goals.

> *“I tend to use ChatGPT more. Because, honestly, if it’s in English. I don’t usually read articles that are too difficult… I tend to put it into AI for summarization first. Because usually the articles are quite long…So I usually get the summary first to see the main points, then decide whether to read the full article…Actually, I sometimes ask AI to summarize the key points from each website. Then I read those summaries…or I use Copilot.””.* P44
>
> *“…because I can use ChatGPT. I’ll ask it to explain something, and if it’s still hard to understand, I’ll ask it to simplify it further.”* P45
>
> *“I’ve used tools like ChatGPT. I have been taking photos of a letter and ask the app to summarize for me. It is easier to understand and saves more time.”* P39

As it pertains to language translation, in interviews conducted at the beginning of the data collection period (before GenAI), participants reflected on their frustration with using online translation tools such as Google Translate, noting that “*translating it will make it worse*” (P09) or that such tools do not help to “*make sense* [of information] *in my language*” (P04), often because the translation was not “*tailored to be understand* [sic] *in our daily…uh…conversation*”. In contrast, in later stages of data collection, AI was seen by some as helping to address these challenges.

> *“I think technology has really improved—especially translation tools. Language barriers are becoming less of an issue thanks to AI and translation software*”. P38
>
> “*I usually go to Google. But now I use ChatGPT more —it helps translate and explain things I don’t know in English… English can be a barrier. Chinese info helps a lot especially for things that are more complicated*”. P39

### Theme 2: “It’s always a mix of trust and scepticism”

#### Recognised need for evaluative judgement

Amongst the *“jungle”* (P49) of information, participants described encountering substantial inconsistency, with a proliferation of divergent accounts across search engines, official health websites, news media and social media platforms.

> *“There are so many websites and differences, so I just look into it—what the differences are. There were a lot of differences, so many results. It made me confused when using Google because there’s so much variation”.* P23
>
> “*What makes it challenging is the overwhelming amount of information and the uncertainty of which one is actually correct*”. P30

While participants noted discrepancies even between ostensibly credible sources, for example “*between the news and the New South Wales Health website”* (P09), variability was particularly salient in social media contexts. Here, participants often described conflicting advice, where *“one source says it’s okay, another says it’s not”* (P28) and felt that they were exposed to “*a lot of misinformation…sometimes even to the point of exaggerations*” (P09) This awareness extended to the growing visibility of influencers in health information ecosystems and the “*situation where everyone thinks they are professionals*” (P06).

In the variable online information environment, most participants appreciated their role in determining the trustworthiness of health information, with many recognising the constant need to “*make decision* [sic] *what to trust, what not to trust, what to believe”* (P01) and determine “*which one is the…um…trusted source”* (P08).

> *“So you still need to evaluate the info yourself. It’s not about blindly following everything”.* P37
>
> *“But sometimes some of these people, these celebrity influencers and stuff will have crazy claims. So you do have to verify them like even, though they have influence and their celebrities, and they’re seen as credible. You still question some of the stuff.”* P36

While two participants expressed that they did not “*try to check*” (P02) whether information is true, others described engaging in active evaluative work as they attempted to “*figure out which one* [information source] *was more accurate*” (P23), reflexively interrogating information by asking questions such as, “*Is this information…true and does it apply to us?*” (P46). Such tasks were “*not easy*” (P34).

> *“You had to go through all the information until you actually identify the one that is related to you or just closely associated with what you’re looking at.”* P52

Here, participants rarely described trust as absolute. Rather, accounts often reflected an ongoing tension between trust and doubt; instead of fully accepting information as authoritative, most participants described maintaining a degree of scepticism, tending not to “*fully trust”* (P28) sources or “*100% agree with what they say*” (P08).

> “*Although I do look at other websites, but as I said, there’s always this doubt in my mind*”. P46

#### Epistemic filtering and the assessment of trustworthiness

Throughout our interviews, participants spoke about how they tried to determine trustworthiness in online spaces. Some participants applied platform-specific heuristics to guide their engagement with online health information, using broad assumptions about the credibility of different platforms to make quick judgements. Functioning as cognitive shortcuts, these heuristics enabled participants to navigate the vast information landscape without needing to verify every individual source.

> “*WhatsApp people like that always like make up something*” P07
>
> *“YouTube is not trustworthy…”* P02

More often, however, determining trust and trustworthiness was a more dynamic and contingent process, constantly being defined and redefined through interactions between the user, the information platform and specific accounts, authors and content creators. Trust was articulated as provisional and comparative, shaped by weighing sources against one another and maintaining a degree of scepticism even toward preferred platforms or creators.

> *On TikTok, I already know which creators I trust….Even though I don’t fully trust them because they’re probably paid, I still follow them. I feel like I need to do my own research first”.* P23

In trying to determine trustworthiness, our participants often described relying on surface-level cues and social proof indicators, as outlined in Table 2.

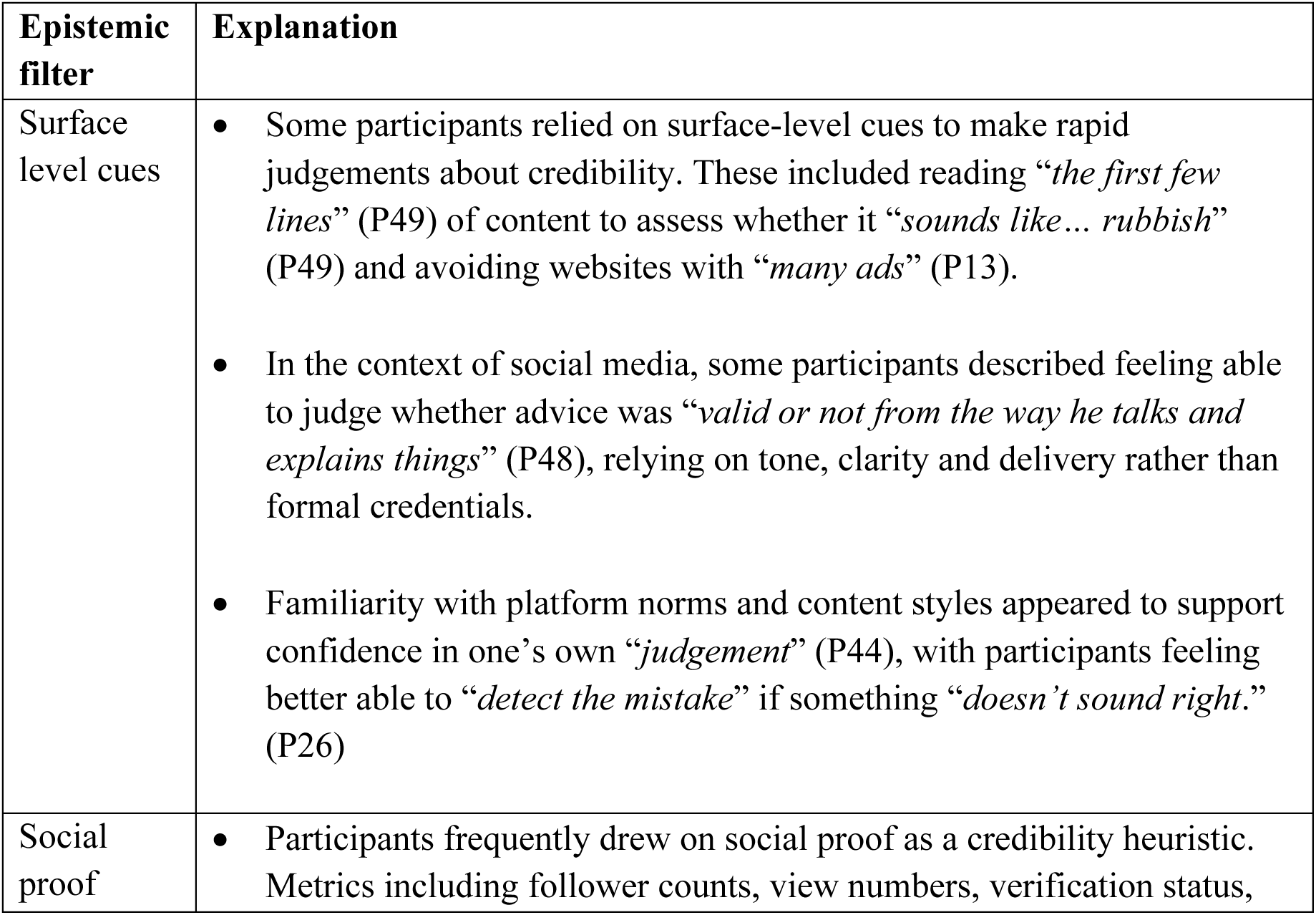
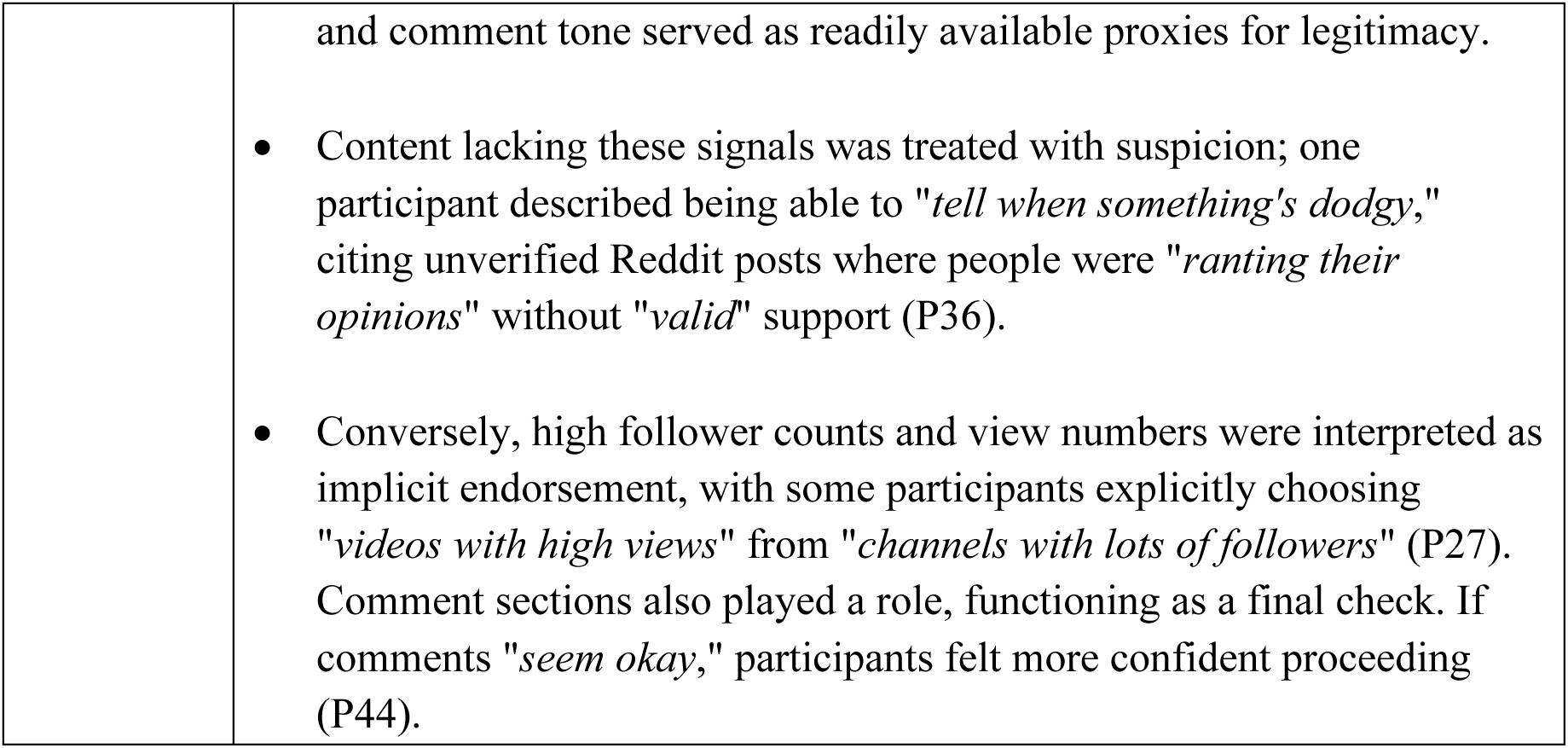

| Epistemic filter | Explanation |
| --- | --- |
| Surface level cues | • Some participants relied on surface-level cues to make rapid judgements about credibility. These included reading *“the first few lines”* (P49) of content to assess whether it *“sounds like... rubbish”* (P49) and avoiding websites with *“many ads”* (P13).<br>• In the context of social media, some participants described feeling able to judge whether advice was *“valid or not from the way he talks and explains things”* (P48), relying on tone, clarity and delivery rather than formal credentials.<br>• Familiarity with platform norms and content styles appeared to support confidence in one's own *“judgement”* (P44), with participants feeling better able to *“detect the mistake”* if something *“doesn't sound right.”* (P26) |
| Social proof | • Participants frequently drew on social proof as a credibility heuristic. Metrics including follower counts, view numbers, verification status, |
|  | and comment tone served as readily available proxies for legitimacy.<br>• Content lacking these signals was treated with suspicion; one participant described being able to *"tell when something's dodgy,"* citing unverified Reddit posts where people were *"ranting their opinions"* without *"valid"* support (P36).<br>• Conversely, high follower counts and view numbers were interpreted as implicit endorsement, with some participants explicitly choosing *"videos with high views"* from *"channels with lots of followers"* (P27). Comment sections also played a role, functioning as a final check. If comments *"seem okay,"* participants felt more confident proceeding (P44). |

Participants also described how they corroborated information across multiple sources before determining what to believe, noting the need to “…*double check it with other sources*” (P09) and “…*check, recheck and recheck*” (P07). Here, participants frequently sought validation from sources positioned as epistemically authoritative, mobilising recognised hierarchies of knowledge and expertise in their efforts to stabilise trust. This included “*official website[s]*” (P01), “*journals*” (P06), and “*authoritative sites to make sure…the news is true*” (P03) as well as hospitals and health professionals in consultations. For some – but not all – participants this included the government.

> *“I’ll look it up first and then go to the doctor. So it depends on the severity. But generally, I prefer to consult a doctor rather than fully trust online information.”* P23
>
> *“I cannot say I could. be, uh, I could trust it 100%. If I had to choose between two things, like, if there was something on the internet and it went against what my doctor was saying, I would go with my doctor.”* P55

#### Professional authority, relatability and the doctor online

While many participants spoke about verifying information with their doctor during in-person consultations, even in online settings, medical authority remained a powerful trust anchor. Many participants described how they “*followed some doctors”* (P28) on social media, engaged with doctors’ content when it surfaced in their online viewing, or recommended doctor’s accounts to others.

> *“If it just comes up randomly when I’m online when I am watching things that a doctor is saying, then I will listen to that information.”* P48
>
> *“Sometimes, you know, there is this doctor explaining about this issue, or something. Yep, on TikTok”.* P31

However, as health information from medical professionals was increasingly encountered in digitally mediated and informal forms, for some participants professional status alone was insufficient to establish epistemic validity. In these contexts, participants recognised that even medical doctors may be *“just trying to get information out for TikTok”* (P48) and needed to draw on additional evaluative strategies to assess the credibility of health professionals.

> *“Oh, yeah, I definitely will go look them online. I’ll see if they have any certification . What their background is. Has anybody who is a known figure advocating them or promoting them, or sort of saying that they’re they’re a reliable source, that they’re valid.”* P36
>
> *“On WeChat , there’s a good account called “Dingxiang Doctor”—very professional and written by real doctors….Xiaohongshu has a lot of scammers. I’ve seen posts in my field that are clearly written by people who don’t understand the topic. So I just browse it casually”.* P37

Participants’ narratives suggest that trust was negotiated through a combination of perceived expertise, communicative style, and levels of social media engagement. That is, surface-level cues and social proof indicators, as described above, were also evident in this context.

> *“If it’s an article, I rarely check who wrote it or open their profile. But if it’s social media, it’s more common—I see a video and wonder, “Is this person legit?” If their profile has lots of followers and a bunch of health-related videos, it feels like—oh, this doctor has real experience. Because nowadays lots of doctors educate through social media”.* P26
>
> *“There are usually several doctors, but most of the time I find one that is more trustworthy than the others The way that he explains things, it would be easier to understand and it sounds more knowledgeable… Because I know that their advice is valid or not from the way he talks and explain things, that’s how I know if its true or not.”* P48

#### AI in epistemic vigilance

Participants described using AI tools both as mechanisms for verification and as objects requiring verification themselves, positioning them ambivalently within their credibility practices. Some reported that they “*check it [health information] with Copilot*” (P53), while others compared AI-generated responses against additional sources to confirm consistency.

> *“… I come to the home and open this video* [from the GP]*. I asked ChatGPT, ‘this is correct?’.” P53*
>
> *“I’ve compared what I get from ChatGPT with other sources, and the information is usually consistent” P45*.

For several participants, AI’s perceived strength lay in its function as an aggregator of multiple sources. Here, credibility was inferred from presumed synthesis; i.e., the idea that AI gathers, consolidates, and surfaces what “most” sources agree upon. In this sense, AI operated as a proxy for consensus. Others emphasised AI’s capacity to pre-process and rank information, describing how it “*reads for you*,” analyses content, highlights popularity and verification markers, and presents a “*comprehensive idea of which one to click on*” (P36). For these participants, AI was seen to reduce the cognitive burden of evaluating competing sources, streamlining comparison and decision-making. Credibility assessment was thus partially outsourced to algorithmic systems that appeared to sort, summarise, and validate information on the user’s behalf.

> *Maybe because it’s not like a direct person who give me the answer, and then, like somehow it gives me more certainty. I don’t know why. I don’t know why, because also, like it derives answer from a bunch of website[s]. And then, if I look at the answer from AI …I feel like it means it’s gathering information. That the most of them* [sic] *have the similar information. And then I feel like they gave me the valid answers. P31*
>
> ***“****It gives you a lot of options on these AI things to compare articles. They sort of reads for you. It analyzes. Or creating clear things for you. It’ll show you. You know what people saying about the article, which one’s more popular, which one’s not. Which one’s verified by experts which one’s not how much it’s been shared. All of that. So, it gives you a very comprehensive idea of which one to click on makes it easier for you. You don’t even have to think much, because it does everything. “* (P36)

However, this trust was not universal. Others expressed scepticism for the very same reason; precisely because AI aggregates information from multiple sources of unknown credibility. For these participants, AI occupied an uneasy middle ground where it was perceived as useful for simplifying complex information, yet fundamentally uncertain as a trustworthy source.

> *“I sometimes use ChatGPT , but since it aggregates from many sources, I don’t fully trust it. I use it more for reference than direct application.” P15*
>
> *“Well, I feel like, well, in my experience I always I would like to suggest ChatGPT when you ask when you have difficulties to gather your thoughts and questions, but then, at the same time, I still have a doubt when using ChatGPT, you know, because I don’t know if this informations[sic] are valid or not. But I feel like it’s the easiest way to have your questions answered.” P31*
>
> *“Yeah, but, like, sometimes I would also try to, like go to the AIs, but, I mean, they’re not really that trustworthy.” P14*

## 4. Discussion

This qualitative study examined how culturally and linguistically diverse adults in Australia access and appraise online health information, foregrounding the individual, social, and cultural resources they mobilise in doing so. To our knowledge, it is the first study to apply a strengths-based, participatory lens to critical health literacy in this population. Our findings speak to both the broad inequities in how digital health information is produced, curated, and distributed in online settings, and to the capacities of communities to critically engage with that information. Our qualitative data attests that culturally and linguistically diverse communities are required to navigate demanding information environments characterised by an overwhelming volume of online health content, much of it not tailored to diverse language needs or literacy levels and drawn from sources that are often difficult to verify (43). Notwithstanding these structural challenges, our strengths-based analysis identified a repertoire of practices through which participants worked to locate understandable information and distinguish credible sources from misinformation. AI tools emerged as a resource in this process, valued for their capacity to simplify and synthesise complex information. However, uncertainty about the credibility of AI itself remained a challenge. These findings extend existing critical health literacy frameworks by demonstrating how individuals draw upon and mobilise diverse epistemic resources to evaluate, interpret, and act on health information in a rapidly evolving digital landscape.

The findings from the current study are broadly consistent with, and extend, existing research on online health information seeking among culturally and linguistically diverse communities. Participants accessed a wide range of platforms including social media, government websites, AI tools, and language-specific applications, reflecting patterns of health information seeking documented in previous studies (44, 45). Our findings also align with a growing body of literature that suggests online health information as easy to locate but overwhelming in volume and complexity, imposing a high cognitive burden and frequently generating confusion (45–48). Where our study extends this literature is in its explicit attention to the strategies participants employed in response to these challenges, and the resources they drew upon to do so. The strategies identified through our interviews reflect Yosso’s (32) linguistic (the intellectual and social skills attained through communication experiences in more than one language and/or style) and navigational capital (skills of manoeuvring through social institutions); social and familial capital were also present and are explored in a supplementary article (Muscat et al., *in development*). For example, participants actively modified search strategies to reduce cognitive burden, sought out simplified formats, and searched in their preferred language to find and clarify information. This extends findings from previous research in which participants similarly refined their approach to information seeking in response to complexity and uncertainty over time (49).

While scepticism toward online health information, particularly on social media, has been documented previously (44), our findings demonstrate that participants actively engage in a range of strategies in attempts to assess the credibility of sources; forms of epistemic vigilance that included cross-referencing information across multiple sources (45), seeking social validation, and anchoring trust in healthcare professionals (29, 50). Our study also identified that trust in social media content was shaped by surface-level cues and social proof, such as follower counts, view numbers, and the tone of user comments. These cues function as cognitive heuristics that enable rapid judgements about credibility under conditions of uncertainty or information overload. Psychological research has shown that individuals frequently rely on such peripheral cues when evaluating information online, often inferring trustworthiness, expertise, or accuracy from indicators of popularity and social endorsement (51). While these heuristics can reduce the cognitive burden associated with evaluating large volumes of information, they may also lead to systematic biases, as signals of engagement are not necessarily aligned with information quality or scientific validity. These findings suggest that efforts to support informed decision-making particularly, but not limited to culturally and linguistically diverse communities, should build on the evaluative strategies people already employ while also providing resources and guidance to strengthen their ability to identify credible information in increasingly complex digital environments.

Our study also provides novel insights into the role of artificial intelligence in health information seeking and evaluation. Previous research has found that the use of AI for obtaining health-related information was more common among people born in non-English-speaking countries, those who spoke a language other than English at home, and those with limited or marginal health literacy (52). Our findings help explain why. Participants described using AI tools as adaptive supports that extended their cognitive and linguistic capabilities, enabling them to translate, simplify, summarise and interrogate health information in ways that aligned with their personal information needs and comprehension goals. At the same time, participants positioned AI ambivalently within their credibility practices, using AI both as a mechanism for verification and as an object requiring verification itself. These findings suggest that AI-mediated health information seeking is shaped by both the quality of the underlying online information environment and individuals’ capacity to engage effectively with AI tools, contributing to emerging debates about AI, trust, and epistemic vigilance in health communication. They position AI literacy as an important extension of health literacy, and highlight the need for further research and co-designed interventions to support critical appraisal in AI-supported health information seeking.

## Strengths and limitations

A key strength of this study is its qualitative design with a large and diverse sample, which enabled in-depth exploration of the experiences of culturally and linguistically diverse adults navigating online health information. The language backgrounds of participants reflect meaningful alignment with the broader Australian population: according to the 2021 Australian Bureau of Statistics Census (53), the five most common languages spoken at home other than English were Mandarin, Arabic, Vietnamese, Cantonese, and Punjabi, which were represented by 38% of our participant sample. We acknowledge, however, the interpretive risks of aggregating data across a culturally diverse sample, potentially overgeneralising the needs of distinct migrant groups (54, 55). The sample was also predominantly female (75%) and highly educated, with 62% holding a Bachelor’s degree or higher; approximately twice the national average among Australians aged 15–74 (56). Given established associations between educational attainment and health literacy, this may overestimate the critical health literacy and digital navigation capabilities of the broader culturally and linguistically diverse population. The underrepresentation of men is also notable, though consistent with wider patterns showing women engage in online health information seeking at higher rates than men (57) despite males showing slightly greater internet usage than women worldwide (58).

The integration of bilingual community co-researchers across all stages of the study including recruitment, data collection, and analysis represents a further significant strength (59), wholly aligned with (inter)national guidelines for engaging consumers and community in research (e.g. (60)). This approach substantially reduced language-based exclusion, enabling participation in preferred languages and facilitating access to communities that research has historically struggled to engage. Bilingual co-researchers also brought cultural knowledge and contextual understanding that enhanced interpretation of participant experiences and helped surface nuances that might otherwise have been missed during framework analysis. However, our methodological approach allowed us to explore only conscious, deliberative decision-making processes—that is, the factors participants reported using and the influences they perceived as shaping their decisions, rather than the often unconscious factors that may operate when searching for information in practice. Consequently, we cannot determine whether decisions made in real-world settings would differ from those described during interviews. For example, early research on online health information seeking found discrepancies between what participants reported in focus groups about how they assessed website credibility and what observational studies revealed about their actual information-seeking behaviours in real time (61).

## Conclusion

These findings point to several priorities for future research and practice. First, while resources to support critical appraisal of online health information exist, few are available in translated forms, culturally adapted for diverse communities, or grounded in a strengths-based approach (Muscat et al., *under review*). As a continuation of this Participatory Action Research, the data generated through this study will inform a co-design process with culturally and linguistically diverse communities to develop resources that are not only linguistically accessible, but that build on the existing skills, networks, and cultural knowledge communities already bring to health information appraisal. Ultimately, advancing critical health literacy requires moving beyond documenting what communities lack toward understanding and strengthening what they already do. By recognising culturally and linguistically diverse communities as active navigators, evaluators, and producers of health knowledge, future research and practice can better support equitable participation in increasingly complex information environments.

## Data Availability

Data are not available

## Acknowledgements

We would like to acknowledge Alicia Wood and Nicola Motton-Palmer from the WSLHD Consumer Council for facilitating consumer partnerships across Western Sydney Local Health District. Your support in coordinating consumer involvement and enabling effective partnership working has been instrumental to the success of this project.

